# Drivers of Frailty from Adulthood into Old Age: Results from a 27-year Longitudinal Population-Based Study in Sweden

**DOI:** 10.1101/19012476

**Authors:** Emma Raymond, Chandra A. Reynolds, Anna K. Dahl Aslan, Deborah Finkel, Malin Ericsson, Sara Hägg, Nancy L. Pedersen, Juulia Jylhävä

## Abstract

**Background:** Frailty is a strong predictor of adverse aging outcomes. However, the longitudinal drivers of frailty are not well understood. This study aimed at investigating the longitudinal trajectories of a frailty index (FI) from adulthood to late life and identifying the predictors of the level and rate of change in FI.

**Methods:** An age-based latent growth curve analysis was performed in the Swedish Adoption/Twin Study of Aging (N=1,842; aged 29-102 years) using data from up to 15 measurement waves across 27 years. A 42-item FI was used to measure frailty at each wave.

**Results:** A bilinear, two-slope model with a turning point at age 65 best described the age-related change in FI, showing that the rate of increase in frailty was more than twice as fast after age 65. Underweight, obesity, female sex, overweight, being separated from one’s co-twin during childhood, smoking, poor social support and low physical activity were associated with a higher level of FI at age 65, with underweight having the largest effect size. When tested as time-varying predictors, underweight and higher social support were associated with a steeper increase in FI before age 65, whereas overweight and obesity were associated with less steep increase in FI after age 65.

**Conclusions:** Predictors for the level and rate of change in frailty are largely actionable and could provide targets for intervention. Underweight increased the risk of higher FI trajectory until age 65, whereas being overweight or obese were associated with slower progression of frailty towards the oldest ages.

## Introduction

Frailty is an age-related condition of sizeable public health importance. It is characterized by increased vulnerability to multiple adverse outcomes, such as disability, hospitalization, and mortality (1). The importance of increased screening and identification of the risk factors among community-dwelling adults has been stressed by recent research (2). However, despite the wealth of research devoted to understanding frailty, its life-course determinants remain poorly understood. Previous research has shown that lifestyle factors in midlife, such as low physical activity (3), sedentary life style (4) and obesity (5) are associated with an increased risk of frailty in old age, but life-course predictors of the rate of change are less well understood and limited to individuals aged 65 years and older (6,7). As recent research by others (8) and us (9-11) suggests that an increase in frailty exhibits a relatively greater mortality risk in younger and middle-aged adults than in old individuals, it is pertinent to identify the determinants of frailty trajectories as early as possible to be able to target the driving forces.

There are several approaches to assess frailty, the Fried frailty phenotype (FP) (12) and the frailty index (FI) developed by Rockwood et al. (13) being the most frequently used ones. The FP views frailty as a physical syndrome and categorizes individuals as robust, pre-frail, and frail based on exhaustion, weight loss, weak grip strength, slow walking speed, and low energy expenditure. The FI is a continuous scale measure that can be constructed from various health-related items, including a minimum of 30 variables that meet the standard criteria (13). Provided that a range of different health domains are included, the FI captures the multidimensional nature of frailty. While both measures predict adverse aging outcomes (14,15), the FI is more sensitive in identifying risks at the lower (healthier) end of the frailty continuum (16,17), and thus is better suited for younger adults.

In the present study, we aimed to add to the understanding of the longitudinal development of frailty and its predictors with a up to 27 years follow-up and 15 waves of repeated measurements in the Swedish Adoption/Twin Study of Aging (SATSA; age range 29-103 years at enrolment). As the development of frailty is a highly multifactorial process and often occurs over a longer period of time and, predictors across various domains were considered and simultaneous changes in key predictors were accounted for.

## Methods

### Sample

SATSA is a longitudinal program in gerontological genetics (18). The participants are same-sexed twins, where some twin pairs have been reared together and some separated before age 11 and reared apart. SATSA was initiated in 1984, ended in 2014 and is comprised of nine questionnaires (Q) and ten in-person testing (IPT) waves. Data collection periods are presented in the Supplement (eFigure 1). New participants in IPT waves were enrolled up until IPT5. SATSA data collection, sampling procedures and the data sets until the seventh Q and seventh IPT have been previously described and are publicly available (19). The present study includes data from 1987 to 2014, from all Q and IPT waves except Q1, IPT1, IPT4 and Q6 (from which an FI could not be constructed), yielding 15 measurement waves. In total, 1,842 individuals (1,074 women, 768 men; 654 complete twin pairs) participated. The number of waves the individuals participated in ranged between one and 15 (mean 5.2, standard deviation [SD] 4.0), where 1,324 individuals (71.9%) participated in three or more waves and 523 (28.4%) participated in seven or more waves.

**Figure 1:**
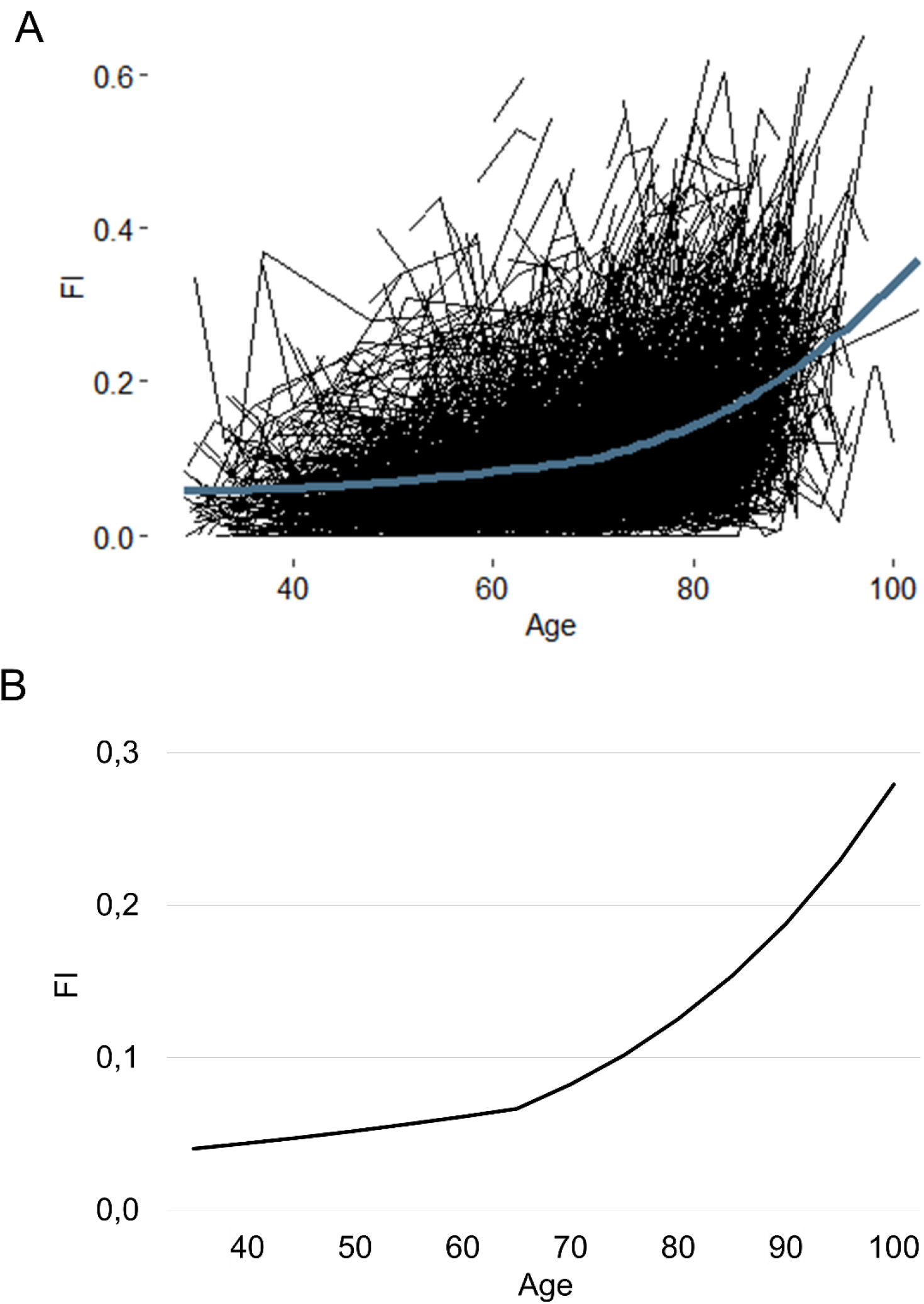
Individual raw trajectories for the frailty index (FI) by age (A) and the estimated population mean FI trajectory from the unconditional bilinear two-slope growth curve model with an inflection point at the age 65 years (B). The blue line in panel A represents a loess-fitted smoothing curve over the data, not assuming within-individual dependency in the observations. Estimates from the unconditional model are based on ln-transformed FI and back-transformed to the original scale to facilitate interpretation (B).

### Study variables

A Rockwood-based FI has previously been created and validated for SATSA(10), and contains 42 self-reported health deficits such as diseases, symptoms, mood and activities in daily living identically constructed across the 15 waves. The FI variables and their scoring are presented in the Supplement (eTable 1). The deficits across the items were scored and the FI was defined as the sum of deficits divided by the number of deficits considered. For example, an individual having 8 deficits had an FI of 8/42=0.19. Although the theoretical maximum of the FI is 1, over 99% of individuals in almost every study cohort have an FI<0.7, indicating that accumulation of deficits beyond this point is lethal (20).

**Table 1.**
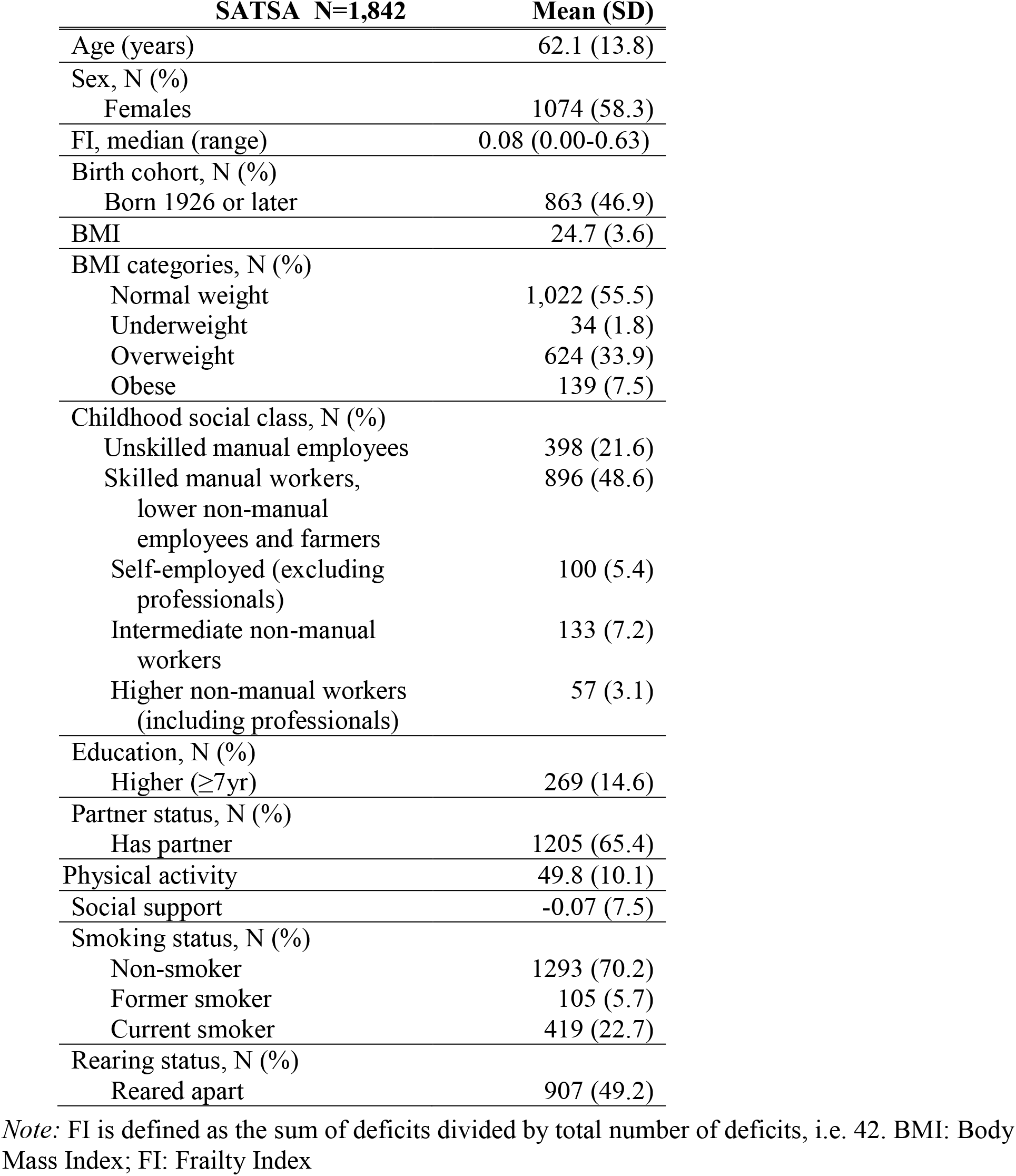
Characteristics of the SATSA participants at baseline (i.e. when the FI was first assessed for each individual). Values are mean (standard deviation, SD) unless otherwise indicated.

Sex was coded as 0=female and 1=male. The birth cohort was coded as 0=born prior 1926 and 1=born 1926 or later, reflecting the division of the old and the middle cohort in the Swedish Twin Registry (STR) (21). Rearing status was coded as 0=reared apart before age 11 (reference) and 1=reared together. Educational attainment and childhood social class were derived from the first questionnaire in 1984 and the STR. Educational attainment was coded as 0=basic education (<7 years; reference) and 1=higher education (≥7 years). Childhood social class, based on self-reported parental occupation for the rearing parents, was coded as 0=unskilled manual employees, 1=skilled manual workers, lower non-manual employees, and farmers, 2=self-employed (excluding professionals), 3=intermediate non-manual workers and 4=higher non-manual workers, including professionals (22). Information about smoking and partner status were derived from the first measurement occasion at which an FI was obtained. Smoking status was coded as 0=non-smoker, 1=former smoker and 2=current smoker, and treated as ordinal in the analysis.. Partner status, which included both marital and non-marital spouse, was coded as 0=no partner (reference) and 1=has partner. Information about body mass index (BMI) was collected at each measurement occasion, calculated as self-reported weight divided by height squared (kg/m^2^), and categorized as follows: 0=normal weight (18.5≤BMI<25.0; reference category), 1=underweight (BMI<18.5), 2=overweight (25.0≤BMI<30.0) and 3=obese (BMI≥30.0). Social support, which was available in Q2, IPT2, Q3, Q4, Q5, Q7, Q8 and Q9, was a standardized continuous score (min −26.8, max 20.6) containing information about the participant’s relationships to friends and relatives (23). A negative score indicated low social support, whilst a positive score indicated high social support. Leisure time physical activity, available in IPT3, Q4, Q5, Q7, Q8 and Q9, was a standardized, continuous score (min 31.4, max 72.4) in which a higher value corresponded to higher physical activity (24).

### Statistical analysis

An age-based latent growth curve model (LGM) was used to assess the longitudinal trajectories in the FI and to identify the determinants for the level and the rate of change in the FI. In LGM, each observation (FI measurement) is a function of a latent intercept, one or several latent slope(s) and random error. The model estimates both a mean trajectory for the entire sample (fixed effects) as well as intra- and interindividual variation around this trajectory (random effects). In this study, variation around the mean trajectory was estimated on three levels. Level 1 constituted the observations. Level 2 and 3 constituted the participants and the twin pairs, respectively. Due to convergence issues, however, we omitted the random slopes for level 3 (twin pairs) and only included a random intercept for level 3. Detailed description of the LGM is provided in the Supplement (eMethods and eFigure 2). Correlations between the FI values across waves were analyzed using Spearman’s rho.

**Figure 2:**
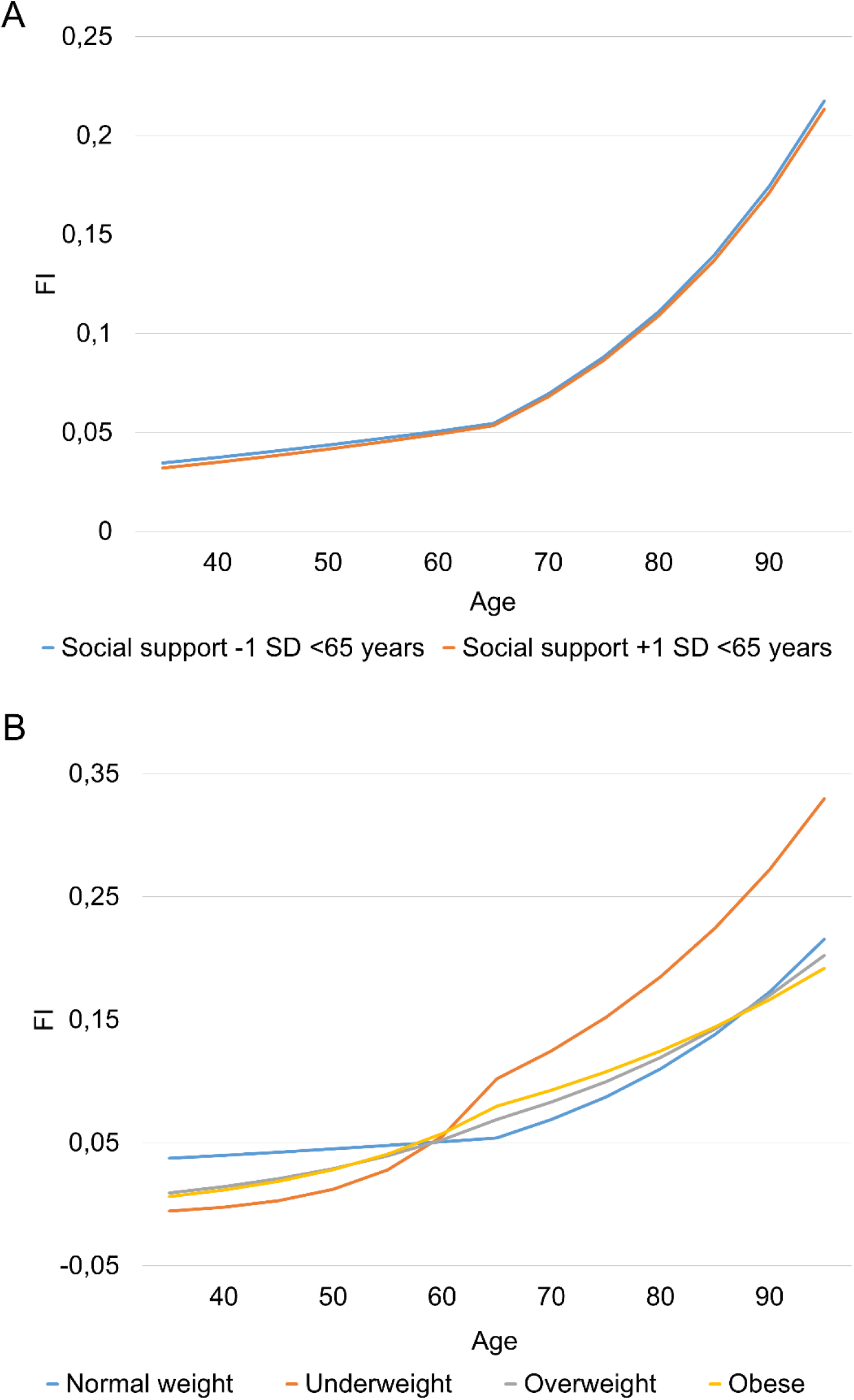
Frailty index (FI) trajectories by repeated time-varying measures on social support (A) and body mass index categories (B) based on a two-slope growth model with an inflection point at age 65 years. For social support, the estimated trajectories are shown for −1 and +1 standard deviation (SD) difference before age 65. The model estimates are based on ln-transformed FI and back-transformed to the original FI scale to facilitate interpretation.

The raw FI trajectory plots were produced in RStudio (R v.3.4.1; Boston, MA: RStudio Inc.) using *ggplot2* (25) (v.3.1.0). To identify the best-fitting functional form (linear, quadratic or bilinear two-slope) to model the change in the FI, age-based unconditional growth curve models were performed in Stata v.15.1 (College Station, TX: StataCorp LP) using the *mixed* command. Best fit was assessed by Akaike Information Criterion (AIC) and Bayesian Information Criterion (BIC) values, where smaller fit statistics indicate better fit.

Sex, birth cohort, rearing status, educational attainment, smoking, childhood social class, partner status, BMI categories, social support and physical activity were tested for their associations with the intercept and the rate of change, which was assessed using a bilinear trajectory with two slopes (see Results). A sensitivity analysis on rearing status was performed to assess whether the degree of relatedness, based on the relationship of the rearing individuals to the twin, is associated with the FI. Information on the degree of relatedness to rearing individuals was available for 1,546/1,842 individuals (83,9%) of whom 935 were in pairs who were reared together, 173 were reared by biological parents (while the co-twin was adopted away), 148 were adopted to relatives and 290 were adopted to non-relatives. Twins reared together were considered as the reference category.

Social support, physical activity and BMI categories were available across several waves (see previously) and hence were additionally tested as time-variant covariates (eMethods and eTable 2). In this model, the associations with both the intercept and slopes were tested using time-varying covariates (social support, physical activity and BMI categories). Smoking status, childhood social class, social support and physical activity were centered at their mean so that the modelled intercepts represent the expected value of FI for an average person in the sample. The BMI categories underweight, overweight and obesity were contrasted to normal weight, as frailty and BMI exhibit a U-shaped association with both high and low BMI increasing the risk of frailty (26,27).

**Table 2:** Latent growth curve models for FI trajectories across 27 years in the SATSA sample. Model A represents the unconditional growth model. Model B tested the associations of all covariates as time-invariant predictors. Model C tested social support, physical activity and BMI categories as time-varying predictors.

| **Association with intercept** | **Model A** | **Model B** | **Model C** |
| --- | --- | --- | --- |
| <sup>a</sup>Intercept | -2.57 (0.02)* | -2.59 (0.04)* | -2.77 (0.05)* |
| Sex |  |  |  |
| Male |  | -0.12 (0.04)* | -0.18 (0.04)* |
| Rearing status |  |  |  |
| Reared apart |  | 0.08 (0.04)* | 0.10 (0.04)* |
| Partner status |  |  |  |
| Has a partner |  | -0.08 (0.04)* |  |
| Smoking status |  | 0.04 (0.02)* | 0.05 (0.02)* |
| Social support |  | -0.008 (0.002)* | -0.009 (0.002)* |
| Physical activity |  | -0.006 (0.001)* | -0.004 (0.001)* |
| BMI categories |  |  |  |
| Underweight |  | 0.43 (0.16)* | 0.56 (0.20)* |
| Overweight |  | 0.09 (0.04)* | 0.21 (0.04)* |
| Obese |  | 0.22 (0.07)* | 0.34 (0.07)* |
| **Association with slope 1 (< 65 years)** |  |  |  |
| Age | 0.014 (0.002)* | 0.013 (0.002)* | 0.010 (0.003)* |
| Social support |  | 0.0007 (0.0002)* | 0.0007 (0.0003)* |
| BMI categories |  |  |  |
| Underweight |  |  | 0.066 (0.031)* |
| Overweight |  |  | 0.005 (0.005) |
| Obese |  |  | 0.015 (0.008) |
| **Association with slope 2 (> 65 years)** |  |  |  |
| Age | 0.038 (0.001)* | 0.038 (0.001)* | 0.042 (0.002)* |
| BMI categories |  |  |  |
| Underweight |  | -0.0005 (0.014) | -0.005 (0.011) |
| Overweight |  | -0.006 (0.003)* | -0.009 (0.004)* |
| Obese |  | -0.002 (0.006) | -0.015 (0.006)* |

*Note:* Estimates (standard error) are based on an ln-transformed FI. Slope 1 represents age<65 years and slope 2 represents age>65 years. Model A has n=1,842 individuals and n=9,534 observations, Model B has n=1,331 individuals and n=8,401 observations, Model C has 1,361 individuals and 3,025 observations.
<sup>a</sup>mean FI at the age of 65 years.
\*p<0.0

Due to the skewed distribution of the FI (left-skewed, data not shown), natural logarithm (ln) of the FI was used in the LGM. Prior to normalization, a constant of 0.01 was added to all FI values to facilitate the transformation of values FI=0. For graphical visualization of the FI trajectories, lnFI values were back-transformed to the original FI scale. Significance level was set to α=0.05.

### Ethics

All participants have given their informed consent. The SATSA study was approved by the Regional Ethics Review Board in Stockholm (Dnr 80:80, 84:61, 86:148, 98:319, 2007/151-31, 2010/657-31/3), 2015/1729-31/5).

## Results

Table 1 provides descriptive information about the study sample at the effective baseline (Q2) where the majority (N=1,477; 80.2%) of the analysis sample had data available. Characteristics of the study sample at each wave are presented in the Supplement (eTable 3). Characteristics of the study sample based on the number of waves the individuals participated in are presented in the Supplement (eTable 4). In comparison to individuals with less than seven complete waves, participants in seven or more waves tended to be younger and more educated, have higher levels of social support, and a lower FI. They were also more likely to have a partner compared to those participating only once or twice. A correlation matrix for the FI across the waves is presented in the Supplement (eTable 5). Most of the correlations were >0.5, and ranged from 0.57 to 0.86 between adjacent waves.

Comparison of the fits of the models based of lnFI (Supplement; eTable 6) indicated that a bilinear two-slope model with an inflection point (intercept) at age 65 years provided the best fit to the data. In this model, the first slope (hereafter denoted as slope 1) represents the change in the FI until the age of 65 and the second slope (hereafter denoted as slope 2) represents the change from this age onwards (Figure 1b). For estimation of the bilinear trajectory, the model used individuals with three or more observations. Individuals with fewer than three data points only contributed to the estimation of the intercept. 1,324 participants (71.9%) participated in at least three waves; thus, they contributed to the estimations of all three latent factors (intercept, slope 1 and slope 2).

The estimated growth curve models using the bilinear fit are presented in Table 2 (only fixed effects are shown). The same models including the random effects are presented in the Supplement (eTable 7). The unconditional (unadjusted) growth model demonstrated that the mean level of lnFI at age 65 years (i.e., the intercept) is −2.57 (corresponding to FI=0.07 on the untransformed scale) and the yearly increase is 0.014 until the age of 65 years and 0.038 after the age of 65 years (Table 2, Model A). Individual raw trajectories for the untransformed FI by age and the estimated bilinear two-slope model for the FI trajectory are presented in Figure 1. The same trajectories but based on the lnFI are presented in the Supplement (eFigure 3).

In Model B (Table 2), the following time-invariant covariates (baseline/first available measurement used) were associated with lower levels of frailty at the age 65: male sex, having a partner, higher social support and higher leisure time physical activity. Being reared apart, underweight, overweight, obese and smoking were associated with higher levels of FI at age 65. Baseline social support and overweight were also associated with the rate of change; higher social support was associated with a steeper increase in FI before age 65 and overweight was associated with slower increase in FI after age 65. Birth cohort was not associated with the FI, and childhood social class and educational attainment were not significant after addition of other covariates so they were excluded from the model. In the sensitivity analysis for the degree of relatedness (based on Model B) where rearing status was replaced with the degree of separation, only those who were adopted to non-relatives had significantly higher FI levels (beta=0.13, standard error [SE]=0.049, p=0.009) compared to those reared together. Twins who stayed with their biological parents while their co-twin was adopted away (beta=0.04, SE=0.058, p=0.5) and twins who were adopted to relatives (beta=0.08, SE=0.06, p=0.2) did not have significantly higher FI levels compared to twins reared together. In Model C that was based on Model B (Table 2), social support, physical activity and BMI categories were tested simultaneously as time-varying covariates. Variables that were associated with the level of FI in this model were akin to Model B, with the exception that partner status was no longer significant and thus excluded from the model. For the associations with the rate of change in FI, higher social support and underweight were associated with a steeper increase in FI before age 65, whereas overweight and obesity were associated with a less steep increase in slope 2 (Table 2, Model C). Physical activity was not associated with the slopes, so its interaction terms with age were excluded from the model. FI trajectories (with lnFI values back-transformed to the original FI scale) based on this model by time-varying measures on social support and BMI categories are presented in Figure 2. Same trajectories but using the lnFI are presented in the Supplement (eFigure 4).

Across all the models A-C, there was significant inter-individual variation in the FI trajectories (Supplement; Random effects in eTable 7). The positive covariance between intercept and slope 1 and negative one between intercept and slope 2 indicted that individuals having a steeper rate of increase up to the age of 65 years have higher FI levels at the inflection point but after that their rate of increase is relatively decreased. The negative covariance between the two slopes, although not statistically significant in Model C, likewise indicated that those having a less steep increase up to the age of 65 years catch up after that. However, random effects for the slopes between twin pairs (level 3) could not be assessed due to convergence issues, so the models only included a random intercept for between-pair effects (Supplement; eTable 7).

## Discussion

This study investigated the developmental trajectories and determinants of FI over an extended period: from adulthood into very old age. The population trajectory indicated a moderate increase up to the age of 65 years – an inflection point after which the increase in FI more than doubled. In a model using baseline measurements for all the predictors, female sex, being reared apart (separated from one’s co-twin in childhood), not having a partner, smoking, low physical activity, underweight, overweight, obesity and poor social support were associated with a higher level of frailty at age 65 (i.e., intercept). Higher baseline social support was also associated with a faster increase in FI before age 65, reflecting only a catch-up growth (convergence) though, as higher social support was associated with a lower FI level at age 65. Baseline overweight was associated with a slower increase in FI after age 65. To address simultaneous changes in the predictors and FI, we extended the model by including time-varying, repeated measurements of social support, physical activity and BMI categories. In this model, higher social support and underweight were associated with a faster rate of increase in the FI before the age of 65 years, and overweight and obesity were associated with a slower rate of increase in the FI after age 65. The decelerated rates of change in FI for those who were obese or overweight were large enough to make the trajectories of FI cross over the FI trajectories for normal weight persons after age 85. Physical activity showed no associations with the rate of change, and having a partner was no longer associated with the level of FI in this model.

Frailty has been traditionally regarded as a syndrome of the old, and longitudinal studies into frailty trajectories including younger individuals or covering the entire adult lifespan are scarce. With repeated measurements of FI up to across 27 years, our study provides new insights into the developmental trajectories of frailty. The fact that we observed a turning point for the FI trajectory at age 65, after which the rate of change more than doubles, suggests that an effective window for prevention might be well before old age. Our estimates for the level and rate of change after age 65 are comparable to those reported in the Longitudinal Aging Study Amsterdam for individuals aged 65+, where they used lnFI in the modeling akin to us (6). However, comparing our (back-transformed) estimates to those for the Swedish population in the Survey of Health, Ageing and Retirement in Europe (SHARE), suggests somewhat lower estimates in our study (28). The SHARE study modeled the rate of change as a single slope for individuals aged 50 and older, which may partly explain the differences. More studies into frailty trajectories covering the entire adult lifespan are nevertheless needed to resolve whether our findings on the bilinear growth and a tipping point at age 65 are generalizable to other populations.

As predictors of frailty have rarely been analyzed in longitudinal settings using repeated measurements both of frailty and the predictors, the results of our study add to the understanding of the independent predictors of frailty. Many of the factors that we found to predict higher levels of frailty, such as female sex, underweight, overweight, obesity, smoking, poor social support, low physical activity and not having a partner have also been identified in previous studies (6,7,26,27). Some studies have also identified low socioeconomic position and lower education associated with higher FI (6,28). In our study, socioeconomic position and education were however not associated with the level of FI after inclusion of other predictors, suggesting that other risk factors may override socioeconomic adversity. As a new risk factor for frailty, we identified being separated from one’s co-twin in childhood to be associated with higher FI levels. Sensitivity analysis for the degree of relatedness to rearing individuals indicated that in comparison to twins reared together, only those who were adopted to non-relatives had significantly higher FI levels, although there was a suggestive trend towards increasing FI levels with increasing degree of relatedness. Socioeconomic adversity alone is unlikely to underlie the association as those non-relative families that could adopt a child at the time were likely wealthier than the families that gave away their child for adoption. The higher FI among the adoptees may thus reflect early life stress, caused by being separated from one’s biological family. A similar finding on childhood stress and frailty was recently reported in the Helsinki Birth Cohort Study; those men who were evacuated abroad unaccompanied by their parents in childhood during World War II had a higher risk of frailty (measured using the FP) in late life compared to non-separated men (29). However, this study found no associations in women. These findings nevertheless call for more research into how different types of childhood stressors may increase the risk of frailty.

Our results are in general in agreement with previous findings on a U-shaped relationship between BMI and the level of frailty (26). Although frailty is partly viewed as a “wasting syndrome” and malnutrition in old age is an established risk factor for it (30), there is a paucity of longitudinal studies looking specifically into underweight and frailty. Our approach using repeated measurements for both FI and BMI allowed us to disentangle this relationship. We not only identified underweight as a major risk factor for higher FI at age 65, but also found an association between time-varying underweight and a faster increase in FI before age 65. The number of underweight individuals in our study was, however, low at baseline and decreased further in the subsequent waves, decreasing our chances to detect potential associations with the rate of change after age 65. The very large effect sizes between underweight and the level and rate of increase in the FI (before age 65) nevertheless indicate that underweight is a key driver of frailty.

Time-varying overweight or obesity were not associated with the rate of change in FI before age 65 but both were associated with a slower increase after age 65. At the age of 85, deceleration rates were large enough to cross over the FI trajectory for normal weight individuals, indicating that older persons being overweight or obese had the fewest frailty deficits. However, as our observations started to get sparse from age 90 onwards, our projections cannot be used to infer whether the decelerated rates would result in significantly lower levels of FI for overweight and obese individuals at the oldest ages. Existence of potential biases, for example due to mortality selection and involuntary weight loss, calls for cautious interpretation. Pertaining to the latter, the category of normal weight (at any given measurement occasion) contains those who are constantly lean as well as those that have lost weight due to disease, making this group potentially heterogeneous in terms of health status. Weight loss, especially in old age, commonly occurs as a result of underlying pathology, which *per se* may be accompanied with increase in frailty. Although less common in old age, high muscle or lean body mass may in turn sometimes manifest as overweight, and being “fat and fit” as opposed to “lean and unfit” has been considered beneficial for some age-related diseases (31-33). Whether such obesity paradox exists for frailty in very old age is unknown. Results from the Helsinki Businessmen Study lend partial support against it (34). They analyzed the association between incident frailty at mean age ∼79 (measured using the FP) and past weight trajectories, and found no protective effect of being constantly overweight or gaining weight (BMI <25 in the beginning and ≥25 in the end) (34). Instead, they found that those who lost weight (BMI ≥25 in the beginning and <25 in the end) had a higher risk of incident frailty compared to constantly normal weight (34). The role of underweight was nevertheless not specifically assessed in this study as none of the men had a BMI <18.5 in midlife.

Interventions based on physical activity have thus far shown most promise in preventing or reversing frailty (35). Although we found that both baseline and time-varying physical activity were associated with lower FI at age 65, the effect size was rather modest and there was no association with the rate of change. One potential explanation is that since this was an observational study, cross-time changes in physical activity are likely to be more subtle than in interventional studies, and a bigger effect size would probably be required to detect significant associations with the rate of change. In addition, physical activity was available only in 6/15 of the measurement occasions, decreasing the number of available repeated measurements. It is also possible that other factors, such as BMI, are stronger in their independent effects, and that there are interactions, such as moderation, between BMI and physical activity. Partially supporting this assertion, a study using latent growth curve analysis on (physical) frailty has shown that obesity has a stronger effect on the progression of frailty in those older individuals who have low physical activity (7).

This study has some limitations. The FI and covariates are based on self-reported data. However, regarding BMI, a previous study in SATSA has demonstrated high correlations (≥0.93) between self-reported and measured BMI and that the misclassification of BMI based on self-reported data is stable over time (36). As pertinent to all studies with long follow-ups into old age, individuals who were younger, less frail and had more favorable values in the covariates provided more measurement occasions, and thus contributed more to the estimates. Moreover, our results cannot be used to infer the direction of the changes between frailty and the predictors. The apparent strengths of this study include the very long follow-up, covering the entire period from adulthood into very old age, ample measurement occasions and availability of a wide array of covariates that enabled us to identify independent predictors of frailty.

In conclusion, our study finds that the predictors for the level of frailty are largely modifiable and could thus be considered as targets for intervention. The association between adoption status and frailty however warrants more research to resolve whether it mostly taps into early life stress or also captures socioeconomic circumstances. As underweight appeared as a driver for increased rate of change before age 65, targeting undernutrition in adulthood might prove useful in preventing future frailty. Any interventional strategy might nevertheless have their effective window before the sharp increase in the rate of change at age 65. Lastly, further studies are needed to resolve whether having extra weight confers some protection against progression of frailty in the advanced ages.

## Data Availability

SATSA questionnaire and in-person testing data up to the seventh testing wave are available at: https://www.icpsr.umich.edu/icpsrweb/NACDA/studies/3843

## Acknowledgements

ER and JJ performed statistical analyses and wrote the manuscript. JJ conceived the study design. CAR contributed to the study design, statistical analysis plan and statistical analyses. ME contributed to the statistical analysis plan. NLP is the founder and principal investigator of the SATSA study. NLP, AKDA, DF, ME and SH contributed to the acquisition of the study variables. All authors contributed to the writing of the manuscript and interpretation of the results.

